# Modulation of premotor cortex response to sequence motor learning during escitalopram-intake

**DOI:** 10.1101/2020.05.19.20105346

**Authors:** Eóin N. Molloy, Karsten Mueller, Nathalie Beinhölzl, Maria Blöchl, Fabian A. Piecha, André Pampel, Christopher J. Steele, Ulrike Scharrer, Gergana Zheleva, Ralf Regenthal, Bernhard Sehm, Vadim V. Nikulin, Harald E. Möller, Arno Villringer, Julia Sacher

## Abstract

The contribution of selective serotonin reuptake inhibitors (SSRIs) to motor learning by inducing motor cortical plasticity remains controversial given diverse findings from positive preclinical data to negative findings in recent clinical trials. To empirically address this translational disparity, we use functional magnetic resonance imaging (fMRI) in a double-blind, randomized controlled study to assess whether 20 mg escitalopram improves sequence-specific motor performance and modulates cortical motor response in 64 healthy female participants. We found decreased left premotor cortex responses during sequence-specific learning performance comparing single dose and steady escitalopram state. Escitalopram plasma-levels negatively correlated with the premotor cortex response. We did not find evidence in support of improved motor performance after a week of escitalopram-intake. These findings do not support the conclusion that one-week escitalopram intake increases motor performance but could reflect early adaptive plasticity with improved neural processing underlying similar task performance when steady peripheral escitalopram levels are reached.

## Introduction

Motor learning is the improved performance of a motor task following practice^1^ and is modulated by monoaminergic transmission in cortical and subcortical motor networks^2,3,4^. Research on this monoaminergic basis of motor learning typically focuses on dopamine signaling in both health^5,6^ and disease^7^. Evidence from rodents^8^ and stroke patients^9^, however, suggests that serotonin also critically modulates motor behavior. Selective serotonin reuptake inhibitors (SSRIs), commonly prescribed medications for depression and anxiety disorders^10^, increase extracellular serotonin and successfully treat post-stroke depression^11^. In the absence of depressive symptoms, several studies have also demonstrated an effect of SSRIs on the recovery of post-stroke motor dysfunction^12^. Notably, the FLAME trial (Fluoxetine for Motor Recovery After Acute Ischemic Stroke^9^) showed approximately 50% motor recovery in 57 patients following combined fluoxetine treatment and physiotherapy, in a multi-center Randomized Controlled Trial (RCT). These findings were further supported by a meta-analysis of 52 RCTs in 4,060 patients, which, however, also acknowledged heterogeneity and methodological shortcomings in a substantial proportion of trials^13^.

Possible mechanisms underlying SSRI modulation of motor performance and learning include anti-inflammatory^14,15^ and neurotrophic effects^16^ such as increased neurogenesis^17^, proliferation^18^, protein expression enhancement^19^, upregulation of beta1-adrenergic receptors^20^, downregulation of GABA-transmission^21,22^, and hippocampal long-term potentiation^23^. These findings suggest that SSRIs may increase responsivity to environmental stimuli, possibly via changes in inhibitory and excitatory balance^24^ and reorganization of cortical networks^25-27^. Studies in humans have provided support for this by demonstrating changes in resting state functional connectivity induced by a single dose of escitalopram^28^. Additionally, decreases in resting state alpha-frequency band induced by tryptophan depletion^29^, which are hypothesized to reflect alterations in the excitatory and inhibitory balance of cortical networks, have been observed in healthy volunteers. Moreover, preliminary functional magnetic resonance imaging (fMRI) evidence has linked decreased functional responses in the motor network with improved motor performance following fluoxetine administration^30-33^.

Recent large-scale RCTs in stroke patients such as the TALOS^34^ and FOCUS trials^35^, involving over 642 and 3,000 patients, respectively, however, do not suggest beneficial effects of SSRIs on functional recovery. Critically, however^36^, these RCTs were conducted against the backdrop of routinely available rehabilitation and did not combine SSRI administration with a clearly defined motor learning paradigm, nor did they assess functional brain responses to SSRI intake. As a result, no previous study, either in healthy participants or in patients, has successfully leveraged prolonged training on an established motor learning paradigm in combination with SSRI-administration and fMRI in an adequately powered sample. Therefore, the hypothesis of whether SSRI administration, specifically in combination with an established motor learning paradigm, induces a beneficial effect on motor learning performance and changes the cortical motor response underlying the learning performance, remains to be tested empirically.

The current study utilizes fMRI to address whether one week of SSRI administration in combination with a sequential motor learning task improves sequence specific motor performance and elicits changes in concurrent cortical motor response during task performance. In a double-blind, randomized controlled pharmaco-fMRI study, we administered 20 mg (to reach 80% serotonin transporter (5-HTT) occupancy)^37^ of escitalopram, the most 5-HTT selective and rapid onset SSRI^38,39^ or placebo, to healthy female participants undergoing parallel fMRI assessment and training on a variant of the sequential pinch force task (SPFT)^40^. Given reports of sex-differences in (i) motor learning^33,41,42^, (ii) fMRI responses during sequential motor control^43^, (iii) motor responses following SSRI-intake^33^, and (iv) sex hormone modulation of serotonin transporter density measures^44,45^ and escitalopram responsivity^46,47^, we chose a healthy and young (to also control for the effects of pathology^48,49^and age^50^) female sample on oral contraceptives, to avoid variance associated with sex and sex hormones on motor learning performance, fMRI response, serotonin transporter density, and SSRI responsivity. Our *a priori* hypotheses were (1) that one week of escitalopram intake would improve sequential motor performance relative to placebo, as assessed by performance in a temporal lag condition on the SPFT, calculated as the time difference between a computer controlled visual stimulus and participant control of a pinch-force device. (2) By specifying this sequence-learning condition in an fMRI contrast (hereafter referred to as the learning contrast, i.e., the difference of functional brain responses between two experimental conditions comprising two levels of task difficulty), we also hypothesized escitalopram-induced changes in fMRI-response in core components of the motor network during task performance.

## Materials and methods

### Participants & eligibility

Eligible individuals were right-handed, aged 18–35 years, with a body mass index (BMI) 18.5-25 kg/m^2^, without history of neurological or psychiatric illness, and female on oral contraceptives for a minimum of three months. Exclusion criteria were medication use, contraindications for MRI, tobacco use, alcohol abuse, positive drug or pregnancy tests, professional musicianship and athleticism, video game use for more than 2 hours, on average, per week, and abnormal QT times in electrocardiogram screenings. In total, 88 participants were screened with 71 enrolled. Analyses included 64 volunteers for the behavioral analysis as 6 (escitalopram=4) chose to discontinue participation and n=1 (placebo) was excluded due to a pre-analytical error in plasma sample acquisition. Sixty volunteers were included in functional imaging analysis as 4 were excluded due to MRI data quality concerns (2 escitalopram) due to head movement. 1 volunteer from the placebo group was excluded due to an artefact in an anatomical sequence and 1 participant from the escitalopram group was excluded due to an artefact detected during acquisition of the functional sequence (Supplementary Figure 1).

### Study design and procedure

The Ethics committee of the Faculty of Medicine, Leipzig University approved all procedures (approval number 390/16-ek), as governed by the Declaration of Helsinki of 2013, and the study was pre-registered at clinicaltrials.gov (ID: NCT03162185). All participants provided written informed consent. Participants were randomized to receive either 20 mg of escitalopram or placebo (mannitol/aerosol) orally for 7 days. Randomization was performed by the Central Pharmacy of Leipzig University with equal condition allocation. Sequential motor training was conducted 5 times (baseline, on day 1 of escitalopram administration (single dose), days 5 and 6 of drug administration, and after 7 days administration – steady state) (Figure 1). Functional magnetic resonance imaging data (fMRI) and serum mature brain-derived neurotrophic factor (mBDNF) samples were acquired at baseline, single dose, and steady state. Electrocardiogram recordings were conducted at single dose, day 4, and steady state to monitor potential changes in QT intervals. Adverse reactions to escitalopram were recorded using the antidepressant side-effects checklist (ASEC)^51^. All participants remained under medical supervision during the experiment. Concentrations of escitalopram in plasma was assessed chromatographically using a quality control sample. Deviation of the measured escitalopram concentration of the sample was tested for an acceptance interval of ±15%. All behavioral and fMRI assessments took place 3 hours after escitalopram or placebo intake to allow for escitalopram to reach maximum levels in serum^52^.

**Figure 1.**
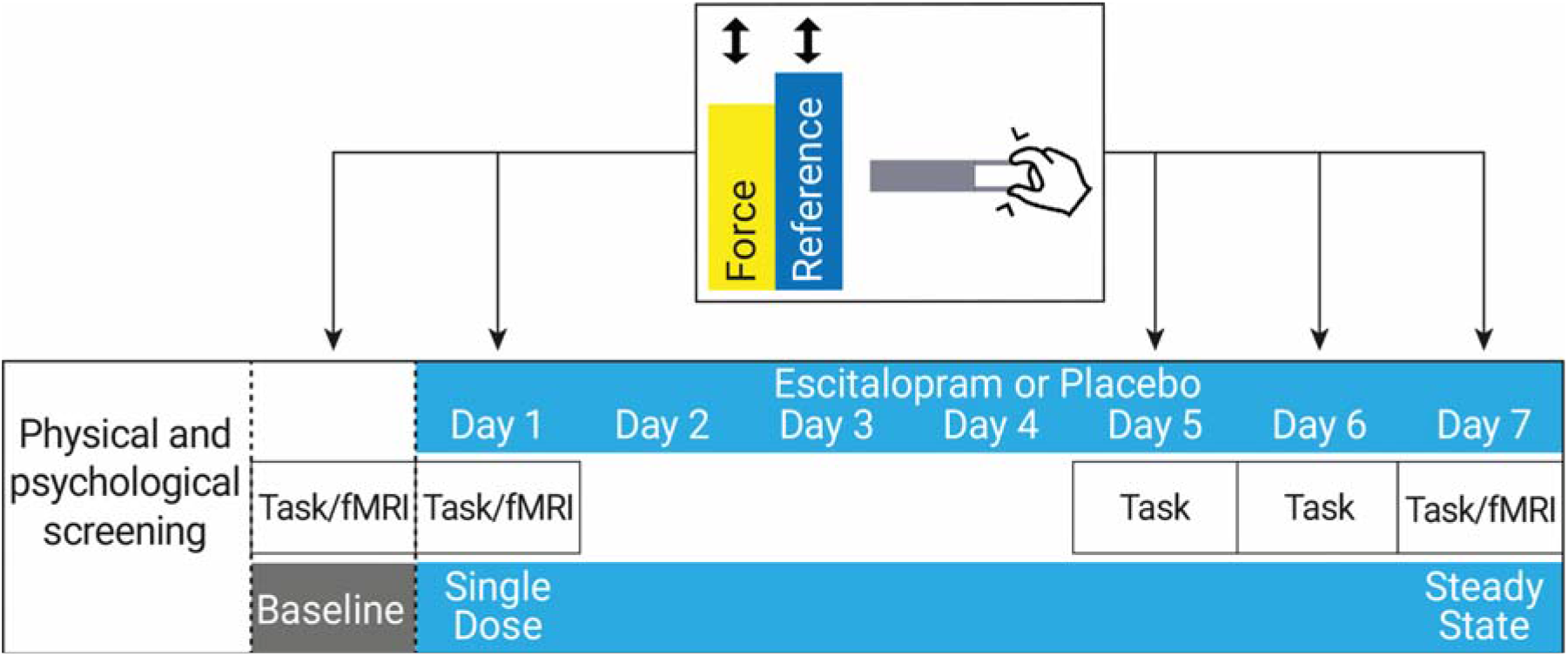
Study design and task: Following baseline, escitalopram or placebo administration took place for 7 consecutive days. Post baseline, motor training took place at single dose (first day), days 5 and 6, and at steady state (day 7). Motor training on days 5 and 6 was completed outside the scanner. fMRI data were acquired at baseline, single dose, and steady state. Task = Sequential Pinch Force Task, fMRI = Functional Magnetic Resonance Imaging, Force = the yellow bar controlled by the participants, the rise and fall of which was required to match the rise and fall of the blue (reference bar, i.e., the bar controlled by a computer).

### Sequential pinch force task

We assessed sequence motor learning using a variant of the sequential pinch force task (SPFT), with Presentation (v16.5) running on WindowsXP. Baseline, single dose, and steady state measurements took place during fMRI, while day 5 and day 6 were conducted outside the scanner on an identical separate device. Task completion involved controlling the rise and fall of a yellow bar (force) via the participant’s thumb and index finger (attenuated to individual strength) while attempting to match the speed of a moving computer controlled blue reference bar (Figure 1). We measured performance in two conditions: (1) a control condition, where the reference bar moves sinusoidally and (2) a sequence-specific learning condition, in which the reference moves in a sequential pattern that remains stable across sessions. A rest condition punctuated training to avoid fatigue. Each session consisted of 5 blocks with 3 trials per block and cycled through simple, rest, and learning. Participants received no feedback regarding performance. To assess performance, we calculated the time difference (lag) in milliseconds between the reference and force bar during the learning trials.

### Demographic data and statistical analysis

Independent samples *t*-tests using the R statistical programming language^53^ tested for potential group differences in age, BMI, downregulated hormonal profile, and on total ASEC scores at single dose and steady state. A power analysis conducted using G*Power^54^ (v.3.1.9.4) assuming statistical power of 95% to detect a significant effect of escitalopram on sequence motor learning *(i.e. learning rate over 5 behavioral assessments compared to placebo)* with a small effect size and an α-level of <0.05 suggested a minimum sample size of 56, with 28 participants per group. To account for potential drop-outs, we aimed to include 60 participants in total.

### Behavioral data preprocessing

All SPFT data were preprocessed using in house MATLAB scripts. Quality control used an outlier labeling approach^55^ implemented in Python (v2.7.15) in which trial, condition, group, and outcome specific interquartile ranges were multiplied by a factor of 1.5 to compute upper and lower bound thresholds.

### Behavioral data analysis

All behavioral data analyses were conducted using R. Normality of data distribution was assessed via visual inspection of Q-Q plots and via a Kolmogorov-Smirnov and Shapiro-Wilk test using the factors of group and time, with the Statistical Package for the Social Sciences (SPSS, version 24).

1. Independent samples *t*-tests assessed baseline group differences using the ‘t.test’ function to assess efficacy of randomization.
2. Comparisons between groups over time employed an omnibus linear random-intercept mixed effect modeling approach using the ‘lmer’ function, within the ‘lme4’ package in R (independent factors: *group, time*, dependent variable: *lag)*. Contributions of each fixed effects were assessed with a likelihood ratio test for improvement of model fit.
3. Post-hoc independent samples *t*-tests were conducted on mean single dose and steady state scores to assess potential group differences at each critical time point of escitalopram-administration. Additionally, the delta (difference between mean performance at steady state compared to baseline) was compared between groups for each outcome via independent samples *t*-tests. Bayes Factor *t*-tests using the ‘ttestBF’ function in the ‘BayesFactor’ package assessed the likelihood of the null hypothesis for all independent sample analyses.
4. Pearson’s correlation analyses assessed potential associations between total ASEC scores and mean lag performance at both single dose and steady state.

### fMRI data acquisition

fMRI data were acquired with gradient-echo echo planar imaging (EPI) on a 3-Tesla MAGNETOM Verio scanner (Siemens, Erlangen, Germany, 32-channel head-coil, flip angle 90°, TR=2000 ms, TE=30 ms, field of view=192×192 mm^2^, 30 slices, 64×64 matrix, 3×3×3mm^3^ 7nominal resolution, 495 volumes, aligned −15° along the anterior to posterior commissure, ~16 minutes). A whole-brain three dimensional T1-weighted Magnetization Prepared Rapid Gradient-echo (MPRAGE) was also acquired at each time point for co-registration^56^ with inversion time, TI=900 ms, TR=2300 ms, TE=2.98 ms, 1×1×1mm^3^ nominal isotropic resolution, ~9 minutes^57^.

### fMRI data analysis – Preprocessing and first-level analysis

Data pre-processing was conducted using SPM12 (v12.7219). Data were realigned, unwarped, normalized to Montréal Neurological Institute (MNI) space, and smoothed with a Gaussian kernel (8mm at full-width-half-maximum). First level analysis was performed for baseline, single dose and steady state separately using a general linear model (GLM) including all three experimental conditions: learning, simple, and rest. In addition, each analysis contained head-movement parameters obtained during preprocessing motion correction. Following parameter estimation, we generated contrast images specific to sequence learning by specifying the learning contrast (i.e., the difference between the learning and simple conditions).

### fMRI data analysis – Group-level analysis

Using contrast images obtained at the first level, whole-brain second level analyses were performed in a stepwise fashion with SPM12 in MATLAB (v9.7). Results were considered statistically significant at a whole-brain cluster-defining threshold of *p*<0.001 corrected at *p*<0.05 using family-wise error (FWE) for multiple comparisons at the whole-brain cluster-level, following which we applied appropriate Bonferroni corrections for the number of contrasts specified.

(1) Following first level analyses, we investigated the learning contrast (i.e. the difference between the learning and the simple condition) for each group at baseline, single dose, and steady state with a one-sample t-test for each group and timepoint independently. Results were Bonferroni corrected for the number of tests performed (n=6).

(2) To assess randomization efficacy, we then conducted an independent-samples t-test to assess potential differences in the learning contrast between groups at baseline.

(3) In order to identify time points of interest within the escitalopram group, we investigated changes in the whole-brain learning contrast across each time point using a one-way repeated measures ANOVA (escitalopram group at three time points – baseline, single dose, and steady state). Here, we assessed all combinations of paired comparisons using Bonferroni correction for the number of tests performed (n=6). Only time points yielding a significant time effect at this corrected threshold were retained for comparison to placebo.

(4) To investigate group differences in the whole-brain learning contrast with respect to results obtained in analyses (3), we specified a flexible factorial model using factors *group* and *time* in which we tested for a *group* by *time* interaction. Subsequent *post-hoc* paired tests were also performed within the same flexible factorial design including Bonferroni correction for the number of possible tests (n=8).

(4a) To ensure that results obtained from our whole-brain flexible factorial analysis (4) were not confounded by intra-subject variance in behavioral motor learning performance, we repeated this analysis with an additional regressor. Here, two behavioral measures for each participant (one for each timepoint) were entered as nuisance covariates in the GLM. For these behavioral measures, we used the behavioral sequence-specific learning measure “lag learning-simple score” (LLSS), as calculated for each participant by subtracting the mean simple condition scores from the mean learning condition lag scores^58^.

(4b) In line with our behavioral analysis, we applied a Bayesian model estimation to investigate the probability of the alternative hypothesis for our interaction contrast. Here, we specified an additional Bayesian model estimation to our flexible factorial design (4) with a medium effect size (equivalent to Cohen’s d = 0.5) and log odds threshold of three, analogous to strong evidence for the alternative hypothesis^59^.

(5) Finally, we tested for correlations within the escitalopram group between (i) motor learning performance and changes in the whole-brain learning contrast and (ii) escitalopram plasma kinetics and changes the in whole-brain learning contrast. We used the LLSS and escitalopram plasma levels, respectively, as a variable of interest within two separate flexible factorial designs. Here, each model was generated using the factors *subject* and *time*, and the variable of interest (i.e. *LLSS* or *plasma escitalopram* levels)^58^. Lastly, we also investigated potential group differences in patterns of brain-behavior correlations by testing an interaction term between the factors *group* and *LLSS*. Results were Bonferroni corrected for multiple comparisons for the number of correlational analyses performed (n=3).

### Analysis of serum mature BDNF levels

A one-way repeated measures ANOVA was implemented in R using the ‘Anova’ function to assess changes in serum mBDNF levels across time. Paired samples *t*-tests in both the escitalopram and placebo groups compared baseline to steady state within each group, separately.

## Results

### Demographics

No differences were observed between groups in any baseline screening measures. Escitalopram levels were within the expected range^52^ (Table 1).

**Table 1.**
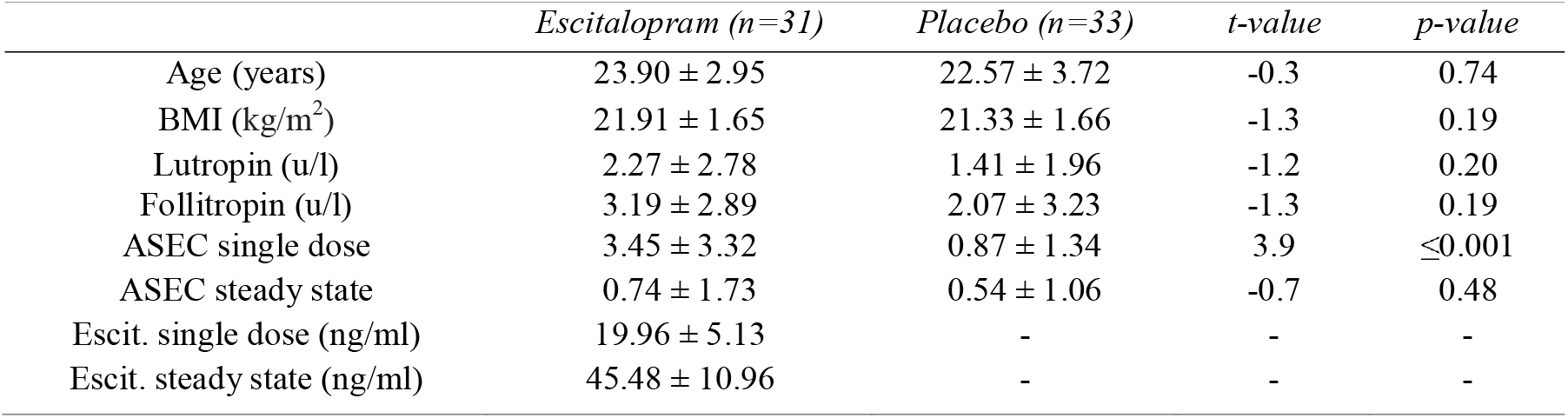
Demographic overview: Group demographic overview and mean single dose and steady state escitalopram plasma concentrations. Group values refer to mean±standard deviation Escit. = escitalopram, ASEC = antidepressant side effect checklist-score, kg/m^2^ = kilogram force per square meter, u/l = units per liter, ng/ml = nanograms/ milliliters.

### Sequence-specific motor learning

Using the Kolmogorov-Smirnov and the Shapiro-Wilk test, we did not observe a significant result for motor training sessions one to four. However, for session 5, we observed a significant result (p=0.003 and p=0.001 for each test, respectively). We did not find any significant differences between the escitalopram and placebo groups on behavioral measures of sequence-specific motor learning:

1. Group comparisons of mean performance at baseline did not show any significant group differences in sequence-specific motor learning behavior (t=-0.25, p=0.80).
2. For group comparisons over time, a mixed effects model including a fixed effect of *time* fit the data significantly better than a random-intercept only model, reflecting a decrease in lag scores (Table 2). The fixed effect of *group* and the interaction of *time* and *group* did not show a significant improvement in fit, demonstrating that, while both groups improved in sequence-specific motor performance over time, they did so comparably (Figure 2).
3. Post-hoc two-sample *t*-tests did not show a significant group difference in mean performance at either single dose or steady state. Comparisons of the delta scores from baseline to steady state did not show any significant differences between groups. Bayes Factor analysis of group comparisons at single dose and steady, as well as the delta, yields moderate evidence in support of the null hypothesis (Table 2).
4. Additionally, correlation analyses did not show an association between total ASEC scores with mean behavioral lag scores at either single dose (r=-0.03, p=0.8,) or at steady state (r=0.11 p=0.37), respectively.

**Figure 2.**
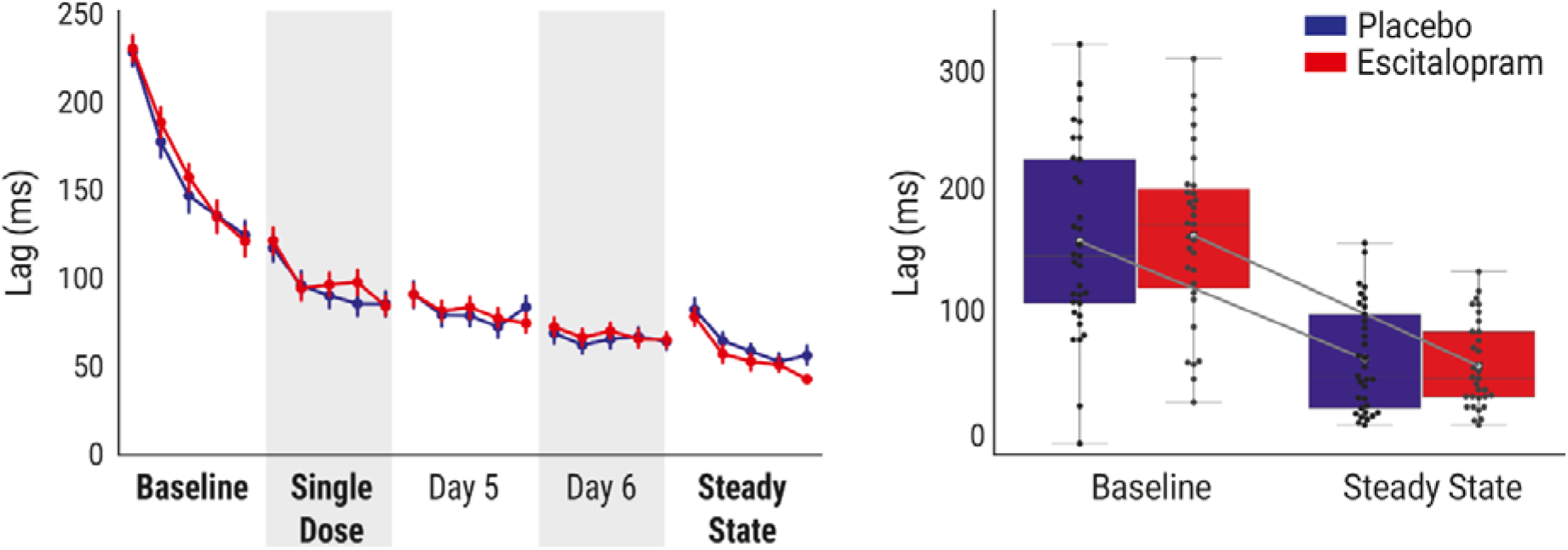
Sequential motor learning: **Left:** Significant improvements in lag scores over 5 days of sequential motor training across both escitalopram and placebo. However, despite a significant learning effect, we observed no significant group differences in performance, nor did we observe an interaction effect. **Right:** Comparison of the rate of change between baseline and steady state yield no significant group differences. Bold fonts indicate training completed in the scanner.

**Table 2.** Comparisons of nested linear mixed effects models and post-hoc testing for sequence-specific lag scores: Model comparisons for computing the omnibus tests for *group* and *time* as well as their interaction effect for both outcome measures. χ2 and respective p-values were computed from a likelihood ratio test between nested models with results of independent samples *t*-tests and corresponding Bayes Factors on mean single dose, steady state, and absolute rate of improvement scores (deltas). BF_01_ = Bayes Factor indicating the likelihood of the alternative hypothesis compared to the null hypothesis.. M±SD = means±standard deviation, LRT = likelihood ratio test, df = degrees of freedom, χ2 = Chi-square. *Significant improvement in model fit.

| *Mixed Effects Modelling* | *Fixed Effects* | *Random effects* | *LRT* |  | *Marginal R<sup>2</sup>* |
| --- | --- | --- | --- | --- | --- |
| | | | $\chi^2$ (df) | *p*-value | |
| Intercept | - | Subject | - | - | 0 |
| Time | time | Subject | 3301.3 (24) | $\leq 0.001^*$ | 0.2917 |
| Group | group+time | Subject | 0.0181 (1) | 0.8931 | 0.2918 |
| Interaction | group*time | Subject | 25.722 (24) | 0.3674 | 0.2934 |
| Post-hoc Testing | Escitalopram (M±SD) | Placebo (M±SD) | *t*-value (df) | *p*-value | Cohen's d/BF<sub>01</sub> |
| Single dose | 99.8±57.9 | 95.1±67.8 | -0.3 (62) | 0.76 | -0.07/0.26 |
| Steady state | 58.1±36.1 | 63.8±44.1 | 0.56 (62) | 0.57 | 0.14/0.29 |
| Delta scores | -107.1±56.6 | -96.7±59.2 | 0.71 (62) | 0.47 | 0.18/0.31 |

### Whole-brain functional MRI responses during sequence-specific motor learning

(1) One-sample *t*-tests across the learning contrast images in each group at each time point show bilateral activation in both the escitalopram and placebo groups at each of baseline, single dose, and steady state (Figure 3).

(2) We did not observe any group differences in the learning contrast fMRI responses at baseline.

(3) Within the escitalopram group, we found a significant decrease of the whole-brain learning contrast in bilateral motor regions when comparing single dose with steady state. We did not observe any significant increases in the learning contrast at any time point.

(4) Comparisons of groups over time reveals decreases in the whole-brain learning contrast in the left premotor cortex of the escitalopram group between single dose and steady state that are not observed in placebo (Figure 3, interaction panel, violet overlay). Post-hoc whole-brain results show a significant decrease in bilateral cortical motor regions in the escitalopram group (Figure 3, escitalopram panel, blue), but not in the placebo group.

(4a) A whole-brain sensitivity analysis controlling for intra-subject variance in task performance replicates this result from analysis (4), showing a significant group by time interaction with a decrease in the learning contrast from single dose to steady state in the escitalopram group in the left premotor cortex (Table 3).

**Table 3.** Escitalopram-induced motor network changes in the learning contrast during sequence motor learning: Results of a whole-brain flexible factorial analysis comparing escitalopram and placebo between single dose and steady state with a *post-hoc* paired comparison (Bonferroni corrected for the number of *post-hoc* tests performed – *α* = 0.006) of single dose and steady state within the escitalopram group only. Results significant at *p*<0.001(unc) cluster threshold corrected with *p*<0.05 Family-wise error on the cluster level. Also shown are the results from a sensitivity analysis with an additional regressor (sensitivity) and a Bayesian model estimation (Bayesian) of the flexible factorial design. p(FWE-corr)= p-value for whole-brain coordinates surviving Family-wise error correction for multiple comparisons, Cluster Extent = number of voxels in cluster. MNI(x,y,z)=MNI peak coordinates, logBF=Log odd threshold, Post-hoc Escit. = escitalopram group post-hoc paired comparisons.

| *Interaction* | *p(FWE-corr)* | *Cluster Extent* | *t-value* | *z-value* | *MNI (x,y,z)* |
| --- | --- | --- | --- | --- | --- |
| Left Premotor Cortex | 0.044 | 82 | 4.36<br>4.17<br>4.10 | 4.04<br>3.88<br>3.82 | -21, 11, 50<br>-15, 8, 59<br>-18, -4, 59 |
| *Interaction (Sensitivity)* |  |  |  |  |  |
| Left Premotor Cortex | 0.017 | 111 | 4.68<br>4.26<br>4.07 | 4.29<br>3.95<br>3.80 | -21, 11, 50<br>-15, 8, 59<br>-27, 5, 56 |
| *Interaction (Bayesian)* | *LogBF Threshold* | *Cluster Extent* | *Cohen's d* |  | *MNI (x,y,z)* |
| Left Premotor Cortex | 3 | 42 | 0.5 |  | -18, 11, 56<br>-18 -4, 56 |
| Post-hoc | *p(FWE-corr)* | *Cluster Extent* | *t-value* | *z-value* | *MNI (x,y,z)* |
| Left Premotor Cortex | <0.001 | 913 | 6.33<br>6.25<br>6.17 | 5.50<br>5.44<br>5.39 | -18, 11, 59<br>-21, 11, 50<br>-27, 8, 59 |
| Right Premotor Cortex | <0.001 | 233 | 6.15<br>5.10<br>4.28 | 5.38<br>4.62<br>3.97 | 30, -4, 41<br>24, -4, 65<br>30, 2, 62 |
| Left Superior Parietal Lobule | <0.001 | 260 | 5.82<br>5.33<br>5.13 | 5.14<br>4.79<br>4.64 | -39, -46, 53<br>-33, -43, 32<br>-36, -46, 41 |
| Left Superior Frontal Gyrus | <0.001 | 295 | 4.87<br>4.53<br>4.31 | 4.44<br>4.17<br>4.00 | -18, 35, 38<br>-18, 44, 20<br>-15, 56, 26 |
| Right Superior Parietal Lobule | <0.001 | 257 | 4.74<br>4.23<br>4.06 | 4.34<br>3.93<br>3.79 | 12, -61, 62<br>18, -61, 50<br>33, -40, 59 |
| Left Postcentral Gyrus | <0.001 | 215 | 4.70<br>4.67<br>3.63 | 4.30<br>4.28<br>3.44 | -9, -46, 74<br>-12, 52, 65<br>-15, -70, 50 |
| Right Middle Temporal Gyrus | 0.002 | 172 | 4.67<br>3.77<br>3.68 | 4.29<br>3.55<br>3.47 | 42, -58, 11<br>33, -58, 23<br>27, -76, 11 |

(4b) Bayesian model estimation yields a significant cluster in the left premotor cortex. Here, we observed strong^59^ evidence in favor of the alternative hypothesis (Table 3).

**Figure 3.**
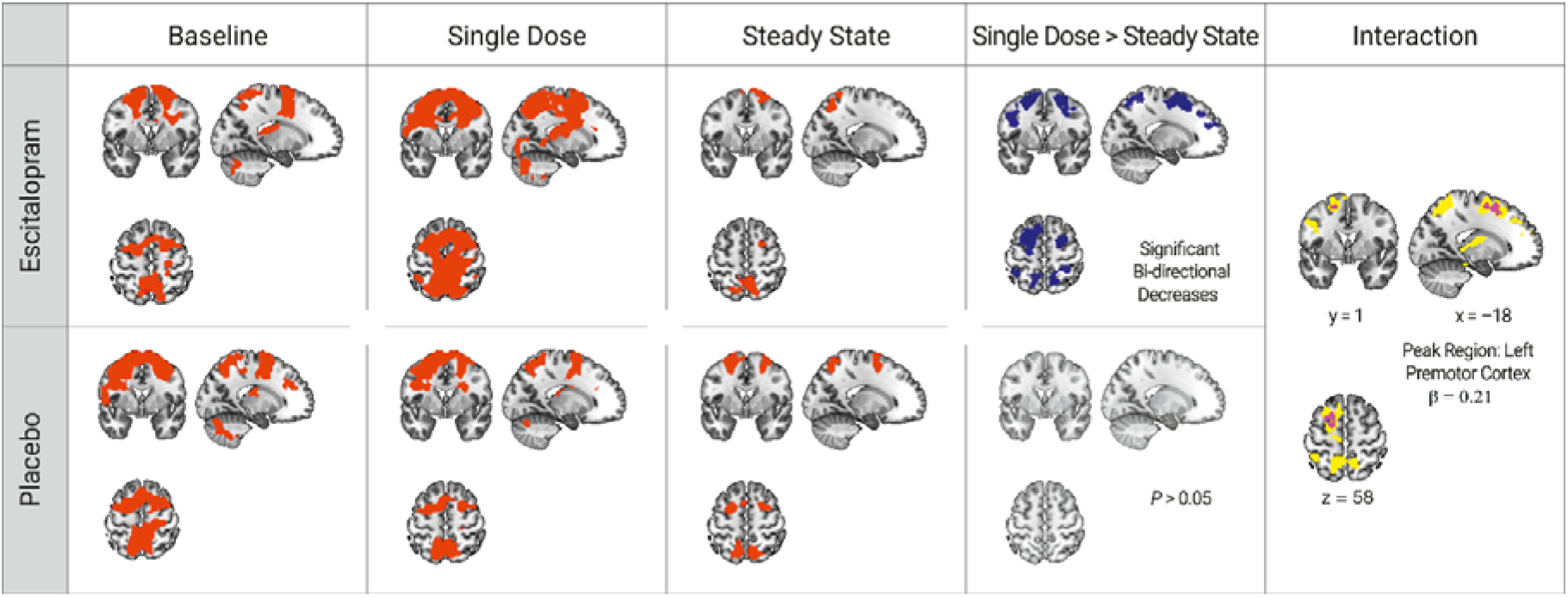
Escitalopram-induced decreases in whole-brain cortical motor responses during sequential motor learning: Orthogonal brain slices showing group dependent changes in the learning contrast over time. Mean functional group response (red) of the escitalopram group (top) and placebo (bottom) at each baseline, single dose, and steady state measurements, as computed by a series of one-sample *t*-tests in SPM12. Single Dose>Steady State: Brain regions in the escitalopram group with significant decreases in the learning contrast (blue) between single dose and steady state (top row) show decreases in bilateral premotor and temporal-parietal regions (Table 3). Comparisons between single dose with steady state in the placebo group do not yield any significant changes across time (bottom). Interaction: Comparisons of groups over time reveal decreases in the whole-brain learning contrast in the left premotor cortex of the escitalopram group that are not observed in placebo (violet). Consideration of behavioral performance as a variable of interest shows brain regions where changes in the learning contrast positively correlate with improvement in motor performance, also with a peak in the left premotor cortex (overlaid in yellow). All results are shown for sequence-specific learning with *p*<0.05 family-wise error (FWE) correction at a cluster forming threshold of *p*<0.001. All orthogonal planes presented are the same. (β = beta value at global maximum coordinate See Supplementary Table 3 for an overview of significant brain regions corresponding to correlation analyses.

(5) Correlation analysis between the change in sequence-specific learning performance with the fMRI signal change in the learning contrast from single dose to steady state within the escitalopram group reveals a significant positive correlation in brain regions including the left premotor cortex (Figure 3; yellow overlay, Supplementary Table 3). However, an interaction contrast testing for differences in the pattern of brain-behavior correlations between groups yields no significant group effect. Correlational analysis between escitalopram plasma levels and the learning contrast in the escitalopram group shows a significant negative correlation, with increases in escitalopram plasma concentration associated with decreases in the learning contrast in bilateral regions including the left premotor cortex and supramarginal gyrus (Figure 4, Supplementary Table 4).

**Figure 4:**
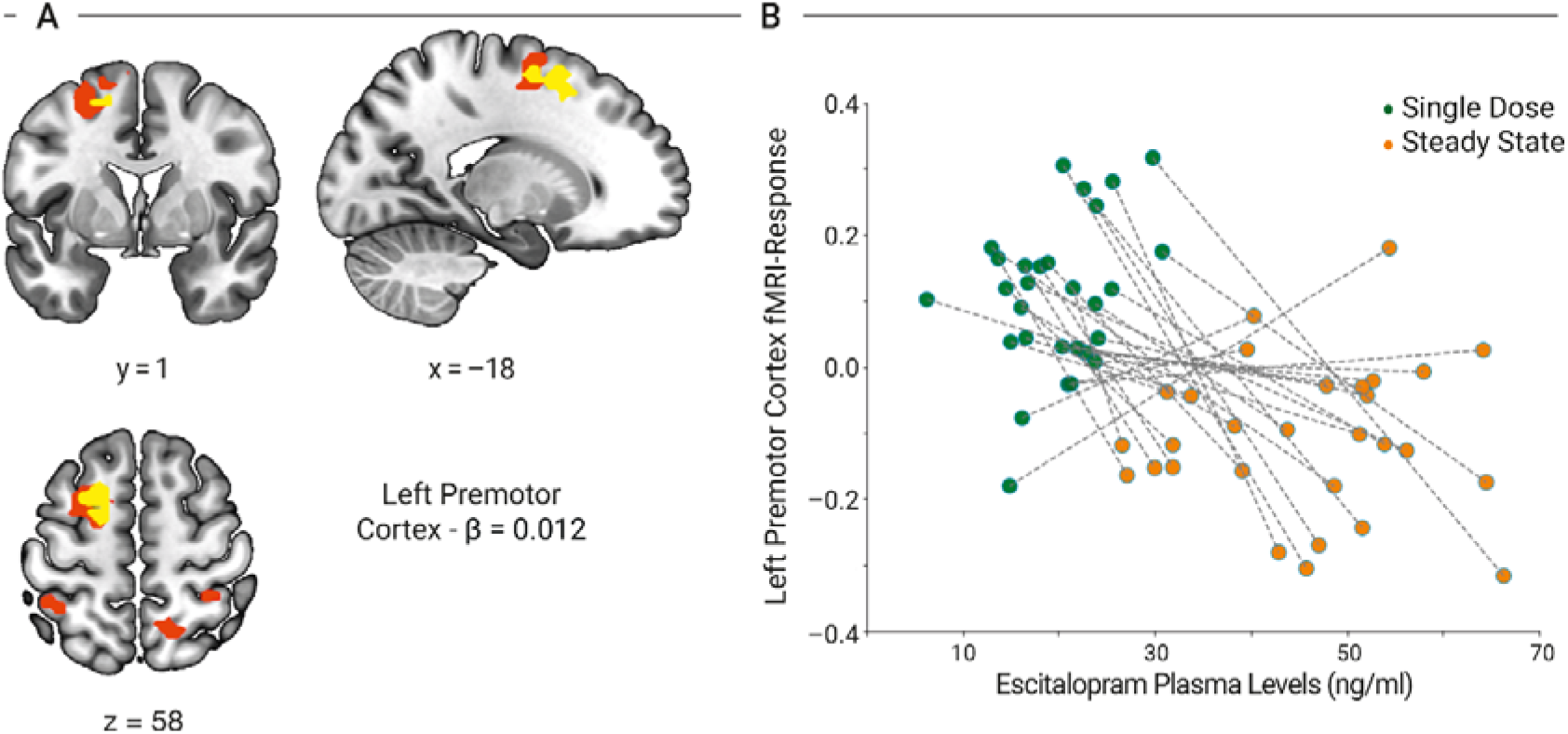
Correlations between escitalopram plasma levels and whole brain cortical premotor response during sequence-specific learning from single dose to steady state: **(A)** Escitalopram plasma concentrations negatively correlate with changes in the whole brain learning contrast in bilateral cortical motor regions, including the premotor cortex (premotor cortex from significant 2×2 interaction overlaid in yellow), with a peak in the left supramarginal gyrus. **(B)** Betas containing parameter estimates for error from the left premotor cortex plotted against escitalopram plasma levels at single dose and steady state, respectively. Results refer to the sequence-specific learning contrast and are shown with *p*<0.05 family-wise error (FWE) correction at a cluster forming threshold of *p*<0.001. Escit. = escitalopram, ng/ml = nanograms/milliliters. β = beta value at premotor MNI coordinates. See Supplementary Table 4 for an overview of significant brain regions.

### Analysis of mature BDNF levels

Analysis of mBDNF levels from baseline to steady state in both groups combined does not reveal any significant changes over time (F(1, 62) = 2.195, *p*=0.12). Paired *t*-tests do not indicate significant changes from baseline to steady state in either the escitalopram (*t* = −1.23, *p*=0.22) or placebo group (*t* =-15, *p*=0.14), respectively.

## Discussion

In this randomized controlled interventional study, we investigated whether administration of 20 mg escitalopram improves motor learning performance and alters functional brain response in the motor network during sequence motor learning. Results show a significant learning effect in sequence-specific motor performance though this rate of improvement does not differ between groups. Additionally, we do not observe any significant group differences at any time point, or in rate of improvement. With a whole-brain fMRI analysis, we find significant escitalopram-induced decreases in the left premotor cortex during sequence-specific learning when comparing single dose and steady state. Moreover, consideration of behavioral performance as a variable of interest during this phase of learning reveals that these changes in the sequence-specific learning contrast positively correlate with improvement in motor performance. Finally, we observe a negative correlation between escitalopram-plasma levels and the fMRI response during the sequence-specific learning contrast in brain regions including the left premotor cortex during task-performance, suggesting a parallel development between escitalopram plasma kinetics and the attenuation of cortical motor response to sequence-specific motor learning.

The lack of an effect of SSRI-administration on motor learning performance differs from previous findings in healthy vounteers^31-33^. These studies, however, were neither powered nor pre-registered to test this as an *a-priori* hypothesis, with six healthy volunteers for five different behavioral tests and one fMRI experiment^31,32^ and nineteen volunteers for six different behavioral assessments^33^. Additionally, we administered escitalopram, and chose a task that may be less cognitively demanding due to repetitive isotonic contractions^60^, possibly creating earlier ceiling effects in healthy adults. These previous findings could be specific to paroxetine, require tasks with more spatial and coordination-oriented sensorimotor components, or may only become apparent after several weeks of administration. Nevertheless, given that we administered the SSRI with the highest transporter selectivity^61^, employed a task that reliably measures sequence motor learning^40^, and tested a sample well-powered to detect robust effect sizes, it is unlikely that this discrepancy is due to the choice of SSRI or motor paradigm alone. Furthermore, our exploratory analysis of mature BDNF levels in plasma did not reveal any significant changes associated with improved motor learning performance in either group. While this is consistent with findings of improved motor performance in healthy volunteers to be unrelated to peripheral BDNF levels^62^, evidence supportive of an association between motor skill learning and increased BDNF levels have also been reported^63^. While future studies should assess potential SSRI modulation of motor learning with additional paradigms, our results do not support a beneficial effect of SSRI-administration on motor learning performance in health.

We do report evidence supportive of our second hypothesis however, with significant decreases in functional responses in left premotor cortex during sequence specific motor learning, relative to placebo (Figure 3). While both increases and decreases in functional brain responses underlie motor learning^64^, this pattern is dependent on differential stages of learning and is defined by multiple parallel processes^65-67^. Early fast learning is accompanied by rapid improvements in performance, followed by slow learning that characterizes a more consolidatory phase^68,69^. Patterns of functional responses observed during this phase are also influenced by the type of task, with explicit learning of repetitive and unchanging sequences hypothesized to lead to faster automation of performance^66,70,71^ and a subsequent reduction in cognitive load needed for task completion. Given the predictable repetition of the learning sequence on our task and the timing of our assessments, it is possible that the observed escitalopram-induced decreases in the learning contrast reflect this automation of responses and subsequent consolidation of sequence learning.

Such a neural consolidation process in response to 1 week of escitalopram-intake is consistent with a recent conceptual model of SSRI influences on post-stroke recovery^72^. The authors propose that acute SSRI exposure changes the excitatory and inhibitory balance with increases in excitatory signaling, allowing for the remodeling of cortical pathways^72^. Subsequent SSRI exposure leads to a reset in homeostasis with a heightening of *γ*-aminobutyric acid (GABA) tone^72^, allowing for remodeled pathways to become engrained as task performance continues. Further support for this interpretation stems from studies identifying an inverse relationship between cortical GABA concentrations and functional brain responses^73,74^ and SSRI administration has been shown to increase cortical GABA levels in rodents^75^ and healthy volunteers^76^. Finally, the observation that the escitalopram-induced decrease in the learning contrast is negatively associated with escitalopram kinetics occurs in a timeframe consistent with that typically required for 5-HT_1A_ autoreceptor desensitization^77^, which could also modulate effective enhancement of cortical GABAergic tone^78^. In summary, it is possible that this escitalopram-induced decrease in premotor response in the learning contrast reflects more effective neural task processing, relative to placebo, in a region central to temporally and visually-oriented motor learning and planning^79-82^. This interpretation is consistent with the hypothesis of an SSRI-induced window of experience-dependent plasticity as an attenuator of neural efficiency during performance^25,26^.

An alternative explanation of this finding is a habituation effect of neural responses during repetitive sequence-specific motor learning that may be emphasized by escitalopram administration. While we report a significant three-way interaction for brain, task, and group, this effect is limited to the comparison between a single dose and steady state and in the escitalopram group only, despite the observation that both groups successfully improve performance over time. It is possible that the neural responses during task performance in the placebo group reflects a simpler order effect, whereby neural responses adapt incrementally, rather than via an adaptive plasticity mechanism. Integration of more direct measures of cortical excitation and inhibition can allow for more fine-grained investigations into acute and subacute SSRI-effects.

Nevertheless, there are some limitations to consider when interpreting these results. We acknowledge that the initial strong learning effect may have masked more subtle modulation of performance with escitalopram at a later training session. Though a known limitation of this task, we chose the SPFT for this well-established and reliable learning effect. While performance reaches a ceiling during the fourth and fifth sessions, as described previously^40^, we still observe a considerable change in performance after the administration of the single dose and subsequent training sessions, thus maintaining the falsifiability of our primary hypothesis. Second, our results may not generalize to males or older adults as our sample consists only of females with standardized downregulation of ovarian hormones. This was a deliberate *a-priori* restriction to eliminate confounds such as sex-differences^33^ and fluctuating endogenous sex hormones on environmental learning^83^ and escitalopram responsivity. While future studies in males, naturally cycling females, as well as healthy aged participants and stroke patients are needed, this choice of sample is also critical given that the majority of preclinical studies test only male samples^84^, posing a critical obstacle to successful translation from healthy models to patients. Third, other studies have gradually increased escitalopram doses for pharmaco-fMRI protocols in healthy participants^85^ to minimize adverse effects. We chose a fixed dose of 20 mg to reliably block 80 % of 5-HTT^37^, an approach previously well tolerated^28^. While four participants discontinued protocol because of adverse effects in the escitalopram group, this was also the case for two placebo participants, and there was no group difference in self-reported side effects at steady state. Finally, fMRI provides an indirect measure of neural activity, which is susceptible to non-neural changes such as vascular uncoupling. Given the functional specificity of the premotor cortex, it is unlikely that these findings are solely driven by changes in global blood flow. We cannot, however, identify underlying molecular mechanisms, which require quantitative measures such as MR-spectroscopy measures of GABA and glutamate or [^11^C]UCB-J positron emission tomography, a recently developed technique for *in vivo* imaging of synaptic plasticity^86,87^.

In conclusion, this is the first study to investigate the effect of steady state escitalopram administration on motor learning in an established sequential motor learning paradigm and the associated brain response in a sufficiently powered sample. In this pre-registered, randomized, controlled, interventional study, we do not find evidence in support of improved performance in response to one week of escitalopram-intake during sequence-specific motor training. A major difference we observe between groups is a decrease in premotor cortical responses during sequence-specific learning performance comparing single dose and steady drug state. Considering previous findings on sequential motor learning and associated neural correlates in the motor network, less premotor response during similar performance may suggest more effective neural processing and greater consolidation of performance^69^. By combining escitalopram administration and sequence-specific motor training for one week, we provide the first empirically tested framework for assessing SSRI-effects on human adaptive motor plasticity in health. Our findings go beyond those of previous studies by employing a well-powered sample with combined longitudinal escitalopram-administration and motor training, in a pre-registered design specifically tailored to empirically test the conceptual network hypothesis of SSRI action^25^. These findings, therefore, address key methodological differences from previous studies in health and in stroke, while simultaneously providing an important step toward understanding the effects of SSRIs on human neural processing during sequence motor learning.

## Data Availability

NA

## Acknowledgements

We thank Heike Schmidt-Duderstedt and Kerstin Flake for assisting with preparation of the figures, Dr. Kristin Ihle for her assistance with medical supervision of all participants during the experiments, and PD Dr. Veronica Witte for statistical discussion.

## Authors’ Contributions Statement

**ENM:** Conceptualization, Investigation, Methodology, Formal analysis, Writing – original draft, Writing – review & editing, Visualization, Funding acquisition

**KM:** Conceptualization, Methodology, Formal analysis, Writing – review & editing, Visualization

**NB:** Conceptualization, Methodology

**MB:** Formal analysis, Writing – review & editing

**FAP:** Formal analysis, Writing – review & editing

**AP:** Methodology

**CJ:** Methodology, Formal analysis

**US:** Conceptualization, Methodology

**GZ:** Conceptualization, Methodology

**RR:** Formal analysis, Writing – review & editing

**BS:** Writing – review & editing

**VVN:** Writing – review & editing

**HEM:** Methodology, Writing – review & editing

**AV:** Writing – review & editing

**JS:**: Conceptualization, Investigation, Methodology, Writing – review & editing, Visualization, Funding acquisition

## References

1. Diedrichsen, J., Kornysheva, K. Motor skill learning between selection and execution. Trends Cogn Sci. 2015; 19(4):227–33. doi: 10.1016/j.tics.2015.02.003.

2. Molina-Luna K., Pekanovic, A., Röhrich, S. et al. Dopamine in motor cortex is necessary for skill learning and synaptic plasticity. PLoS One. 2009; 17;4(9):e7082. doi:10.1371/journal.pone.0007082.

3. Rioult-Pedotti, MS., Pekanovic, A., Atiemo, CO et al. Dopamine Promotes Motor Cortex Plasticity and Motor Skill Learning via PLC Activation. PLoS One. 2015; 10(5):e0124986. doi: 10.1371/journal.pone.0124986.

4. Vitrac, C., Benoit-Marand, M. Monoaminergic Modulation of Motor Cortex Function. Front. Neural Circuits 2017; 11:72. doi: 10.3389/fncir.2017.00072.

5. Flöel, A., Breitenstein, C., Hummel, F. et al. Dopaminergic influences on formation of a motor memory. Ann Neurol. 2005; 58(1):121–30. doi:10.1002/ana.20536.

6. Flöel, A., Garraux, G., Xu, B. et al. Levodopa increases memory encoding and dopamine release in the striatum in the elderly. Neurobiol Aging. 2008; 29(2):267–79. doi:10.1016/j.neurobiolaging.2006.10.009

7. Magrinelli F, Picelli A, Tocco P, et al. Pathophysiology of Motor Dysfunction in Parkinson’s Disease as the Rationale for Drug Treatment and Rehabilitation. Parkinsons Dis. 2016; 2016:9832839. doi:10.1155/2016/9832839

8. Maya Vetencourt, JF., Sale, A., Viegi, A. et al. The antidepressant fluoxetine restores plasticity in the adult visual cortex. Science, 2008; 320(5874), 385–388. doi:10.1126/science.1150516.

9. Chollet, F., Tardy, J., Albucher, JF. et al. Fluoxetine for motor recovery after acute ischaemic stroke (FLAME): a randomised placebo-controlled trial. Lancet Neurol, 2011; 10(2), 123–130. doi:10.1016/S1474-4422(10)70314-8.

10. Cipriani A, Furukawa TA, Salanti G, et al. Comparative efficacy and acceptability of 21 antidepressant drugs for the acute treatment of adults with major depressive disorder: a systematic review and network meta-analysis. Lancet. 2018;391(10128):1357–1366. doi:10.1016/S0140-6736(17)32802-7

11. Hackett, ML., Anderson, CS., House, A, et al. Interventions for preventing depression after stroke. Cochrane Database Syst Rev. 2008; 16;(3):CD003689. doi: 10.1002/14651858.CD003689.pub3.

12. Acler, M., Robol, E., Fiaschi, A. et al. A double blind placebo RCT to investigate the effects of serotonergic modulation on brain excitability and motor recovery in stroke patients. Journal of Neurology2009; 256(7):1152–8. doi:10.1007/s00415-009-5093-7.

13. Mead, GE., Hsieh, CF., Hackett, M. Selective serotonin reuptake inhibitors for stroke recovery. JAMA. 2013; 310(10):1066–7. doi: 10.1001/jama.2013.107828.

14. Lim, CM., Kim, SW., Park, JY. et al. Fluoxetine affords robust neuroprotection in the postischemic brain via its anti-inflammatory effect. J Neurosci Res. 2009; 87(4): 1037-45. doi: 10.1002/jnr.21899.

15. Tynan, RJ., Weidenhofer, J., Hinwood, M. et al. A comparative examination of the anti-inflammatory effects of SSRI and SNRI antidepressants on LPS stimulated microglia. Brain Behav Immun. 2012; 26(3):469–79. doi:10.1016/j.bbi.2011.12.011.

16. Schmidt, HD., Duman, RS. The role of neurotrophic factors in adult hippocampal neurogenesis, antidepressant treatments and animal models of depressive-like behavior. Behav Pharmacol. 2007; 18(5-6):391–418. doi: 10.1097/FBP.0b013e3282ee2aa8

17. Malberg, JE., Eisch, AJ., Nestler, EJ. et al. Chronic antidepressant treatment increases neurogenesis in adult rat hippocampus. J Neurosci. 2007; 15;20(24):9104–10. doi: https://doi.org/10.1523/JNEUR0SCI.20-24-09104.2000.

18. Lyons L, ElBeltagy M, Umka J. et al. Fluoxetine reverses the memory impairment and reduction in proliferation and survival of hippocampal cells caused by methotrexate chemotherapy. Psychopharmacology (Berl). 2011;215(1):105–115. doi:10.1007/s00213-010-2122-2

19. Cooke JD, Grover LM, Spangler PR. Venlafaxine treatment stimulates expression of brain-derived neurotrophic factor protein in frontal cortex and inhibits long-term potentiation in hippocampus. Neuroscience. 2009;162(4):1411–1419. doi:10.1016/j.neuroscience.2009.05.037

20. Pälvimäki EP, Laakso A, Kuoppamäki M, et al. Up-regulation of beta 1-adrenergic receptors in rat brain after chronic citalopram and fluoxetine treatments. Psychopharmacology (Berl). 1994;115(4):543–546. doi:10.1007/BF02245579

21. Yan, Z. Regulation of GABAergic inhibition by serotonin signalling in prefrontal cortex. Molecular Neurobiology2002; 26(2-3) 203–216. doi: 10.1385/MN:26:2-3:203

22. Choi HC, Kim YI, Song HK, et al. Effects of selective serotonin reuptake inhibitors on GABAergic inhibition in the hippocampus of normal and pilocarpine induced epileptic rats. Brain Res. 2010;1357:131–141. doi:10.1016/j.brainres.2010.08.010

23. Mlinar, B., Stocca, G., Corradetti, R. Endogenous serotonin facilitates hippocampal long-term potentiation at CA3/CA1 synapses. J Neural Transm(Vienna). 2015; 122(2): 177–85.doi: 10.1007/s00702-014-1246-7.

24. Batsikadze G, Paulus W, Kuo MF, et al. Effect of serotonin on paired associative stimulation-induced plasticity in the human motor cortex. Neuropsychopharmacology. 2013;38(11):2260–2267. doi:10.1038/npp.2013.127

25. Castrén, E. Is mood chemistry? Nat Rev Neurosci, 2005;6(3), 241–246. doi:10.1038/nrn1629

26. Castrén, E. Neuronal network plasticity and recovery from depression. JAMA Psychiatry, 2013; 70(9), 983-989. doi:10.1001/jamapsychiatry.2013.1

27. Siepmann T, Penzlin AI, Kepplinger J, et al. Selective serotonin reuptake inhibitors to improve outcome in acute ischemic stroke: possible mechanisms and clinical evidence. Brain Behav. 2015;5(10):e00373. Published 2015 Sep 23. doi:10.1002/brb3.373

28. Schaefer, A., Burmann, I., Regenthal, R., et al. Serotonergic modulation of intrinsic functional connectivity. Current Biology, 2014; 24(19), 2314–2318. doi: 10.1016/j.cub.2014.08.024.

29. Knott, VJ., Howson, AL., Perugini, M. et al. The Effect of Acute Tryptophan Depletion and Fenfluramine on Quantitative EEG and Mood in Healthy Male Subjects. Biological Psychiatry; 1999; (46) 229–238.

30. Loubinoux, I., Boulanouar, K., Ranjeva, JP. et al. Cerebral functional magnetic resonance imaging activation modulated by a single dose of the monoamine neurotransmission enhancers fluoxetine and fenozolone during hand sensorimotor tasks. J Cereb Blood Flow Metab. 1999; 19(12):1365–75. doi: 10.1097/00004647-199912000-00010

31. Loubinoux, I., Pariente, J., Boulanouar, K. et al. A single dose of the serotonin neurotransmission agonist paroxetine enhances motor output: double-blind, placebo-controlled, fMRI study in healthy subjects. Neuroimage, 2002a; 15(1), 26–36. doi:10.1006/nimg.2001.0957.

32. Loubinoux, I., Pariente. J., Rascol, O. et al. Selective serotonin reuptake inhibitor paroxetine modulates motor behavior through practice. A double-blind, placebo-controlled, multi-dose study in healthy subjects. Neuropsychologia. 2002b; 40(11):1815–21.

33. Loubinoux, I., Tombari, D., Pariente, J. et al. Modulation of behavior and cortical motor activity in healthy subjects by a chronic administration of a serotonin enhancer. Neuroimage. 2005; 27(2):299–313. doi:10.1016/j.neuroimage.2004.12.023

34. Kraglund, KL., Mortensen, JK., Damsbo, AG., et al. Neuroregeneration and Vascular Protection by Citalopram in Acute Ischemic Stroke (TALOS). Stroke. 2018; 49(11):2568–2576. doi: 10.1161/STROKEAHA.117.020067.

35. FOCUS Trial Collaboration. Effects of fluoxetine on functional outcomes after acute stroke (FOCUS): a pragmatic, double-blind, randomised, controlled trial. Lancet. 2019; 393(10168):265–274. doi: 10.1016/S0140-6736(18)32823-X.

36. van der Worp, BH. Fluoxetine and recovery after stroke. Lancet. 2019; 393(10168):206-207. doi: 10.1016/S0140-6736(18)32983-0.

37. Klein N, Sacher J, Geiss-Granadia T, et al. In vivo imaging of serotonin transporter occupancy by means of SPECT and [123I]ADAM in healthy subjects administered different doses of escitalopram or citalopram. Psychopharmacology (Berl). 2006;188(3):263–272. doi:10.1007/s00213-006-0486-0

38. Kasper, S., Spadone C., Verpillat P. et al. Onset of action of escitalopram compared with other antidepressants: results of a pooled analysis. Int Clin Psychopharmacol. 2006; 21(2):105–10.

39. Sanchez, C., Reines, EH., Montgomery, SA. A comparative review of escitalopram, paroxetine, and sertraline: Are they all alike? Int Clin Psychopharmacol. 2014; 29(4): 185–96. doi: 10.1097/YIC.0000000000000023.

40. Gryga M, Taubert M, Dukart J, et al. Bidirectional gray matter changes after complex motor skill learning. Front Syst Neurosci. 2012; 6:37. doi:10.3389/fnsys.2012.00037

41. Moreno-Briseño, P., Díaz, R., Campos-Romo, A. et al. Sex-related differences in motor learning and performance. Behavioral and brain functions: BBF, 2010; 6(1),74. https://doi.org/10.1186/1744-9081-6-74

42. Dorfberger, S., Adi-Japha, E., & Karni, A. Sex differences in motor performance and motor learning in children and adolescents: an increasing male advantage in motor learning and consolidation phase gains. Behavioural brain research, 2009;198(1), 165–171. https://doi-org.browser.cbs.mpg.de/10.1016/j.bbr.2008.10.033

43. Lissek S, Hausmann M, Knossalla F, et al. Sex differences in cortical and subcortical recruitment during simple and complex motor control: an fMRI study. Neuroimage. 2007;37(3):912–926. doi: 10.1016/j.neuroimage.2007.05.037

44. Frokjaer VG, Pinborg A, Holst KK, et al. Role of Serotonin Transporter Changes in Depressive Responses to Sex-Steroid Hormone Manipulation: A Positron Emission Tomography Study. Biol Psychiatry. 2015;78(8):534–543. doi:10.1016/j.biopsych.2015.04.015

45. Kranz GS, Wadsak W, Kaufmann U, et al. High-Dose Testosterone Treatment Increases Serotonin Transporter Binding in Transgender People. Biol Psychiatry. 2015;78(8):525–533. doi:10.1016/j.biopsych.2014.09.010

46. LeGates, T., Kvarta, MD., Thompson, SM. Sex differences in antidepressant efficacy. Neuropsychopharmacology; 2009; 44(1):140–154. doi: 10.1038/s41386-018-0156-z.

47. Sramek, J.J., Murphy, M.F., Cutler, N.R. Sex differences in the psychopharmacological treatment of depression. Dialogues in clinical neuroscience, 2016;18(4), 447–457.

48. Pasqualetti G., Gori G., Blandizzi C. et al. Healthy volunteers and early phases of clinical experimentation. Eur J Clin Pharmacol.; 2010; 66(7):647–53. doi:10.1007/s00228-010-0827-0.

49. Karakunnel JJ, Bui N, Palaniappan L, et al. Reviewing the role of healthy volunteer studies in drug development. J Transl Med. 2018;16(1):336. doi:10.1186/s12967-018-1710-5

50. van Dyck CH, Malison RT, Seibyl JP, et al. Age-related decline in central serotonin transporter availability with [(123)I]beta-CIT SPECT. Neurobiol Aging. 2000;21(4):497–501.doi:10.1016/s0197-4580(00)00152-4

51. Uher R, Farmer A, Henigsberg N, et al. Adverse reactions to antidepressants [published correction appears in Br J Psychiatry. 2010 May;196(5):417]. Br J Psychiatry. 2009;195(3):202–210. doi:10.1192/bjp.bp.108.061960

52. Rao, N. The Clinical Pharmacokinetics of Escitalopram. Clinical Pharmacokinetics 2007; 46(4):281–90. DOI: 10.2165/00003088-200746040-00002.

53. R Core Team. R: A language and environment for statistical computing. R Foundation for Statistical Computing, Vienna, Austria. 2013

54. Faul, F., Erdfelder, E., Lang, AG. et al. G*Power 3: a flexible statistical power analysis program for the social, behavioral, and biomedical sciences. Behav Res Methods. 2007; 39(2):175–91. doi:10.3758/bf03193146.

55. Iglewicz, B., Banerjee, S. A simple univariate outlier identification procedure. Proceedings of the Annual Meeting of the American Statistical Association. 2001.

56. Mugler JP 3rd, Brookeman JR. Three-dimensional magnetization-prepared rapid gradient-echo imaging (3D MP RAGE). Magn Reson Med. 1990;15(1):152–157. doi:10.1002/mrm.1910150117

57. Streitbürger DP, Pampel A, Krueger G, et al. Impact of image acquisition on voxel-based-morphometry investigations of age-related structural brain changes. Neuroimage. 2014;87:170–182. doi:10.1016/j.neuroimage.2013.10.051

58. Holiga Š, Mueller K, Möller HE, et al. Motor matters: tackling heterogeneity of Parkinson’s disease in functional MRI studies. PLoS One. 2013;8(2):e56133. doi:10.1371/journal.pone.0056133

59. Han, H., Park, J. Using SPM 12’s Second-Level Bayesian Inference Procedure for fMRI Analysis: Practical Guidelines for End Users. Front. Neuroinform. 2018;12:1. doi:10.3389/fninf.2018.0000

60. Hardwick, RM., Rottschy, C., Miall, RC. et al. A quantitative meta-analysis and review of motor learning in the human brain. Neuroimage. 2013; 15; 67:283–97. doi: 10.1016/j.neuroimage.2012.11.020.

61. Owens, MJ., Knight, DL., Nemeroff, CB. Second-generation SSRIs: human monoamine transporter binding profile of escitalopram and R-fluoxetine. Biol Psychiatry. 2001; 50(5):345–50. doi: 10.1016/s0006-3223(01)01145-3

62. Baird JF, Gaughan ME, Saffer HM, et al. The effect of energy-matched exercise intensity on brain-derived neurotrophic factor and motor learning. Neurobiol Learn Mem. 2018;156:33–44. doi:10.1016/j.nlm.2018.10.008

63. Grégoire CA, Berryman N, St-Onge F, et al. Gross Motor Skills Training Leads to Increased Brain-Derived Neurotrophic Factor Levels in Healthy Older Adults: A Pilot Study. Front Physiol. 2019;10:410. Published 2019 Apr 12. doi:10.3389/fphys.2019.00410

64. Wymbs, FN., Grafton, ST. The Human Motor System Supports Sequence-Specific Representations over Multiple Training-Dependent Timescales. Cerebral Cortex, 2015; 25(11) 4213–4225, doi: https://doi.org/10.1093/cercor/bhu144

65. Penhune, VB., Steele, CJ. Parallel contributions of cerebellar, striatal and M1 mechanisms to motor sequence learning. Behav Brain Res. 2012; 15;226(2):579–91. doi:10.1016/j.bbr.2011.09.044.

66. Toni, I., Krams, M., Turner, R. et al. The time course of changes during motor sequence learning: a whole-brain fMRI study. Neuroimage. 1998; 8(1):50–61. doi: https://doi.org/10.1006/nimg.1998.0349

67. Luft, AR., Buitrago, MM. Stages of motor skill learning. Mol Neurobiol 2005;32(3):205–16. doi:10.1385/MN:32:3:205.

68. Karni A, Meyer G, Rey-Hipolito C, et al. The acquisition of skilled motor performance: fast and slow experience-driven changes in primary motor cortex. Proc Natl Acad Sci USA. 1998;95(3):861–868. doi:10.1073/pnas.95.3.861

69. Hotermans, C., Peigneux, P., Maertens de Noordhout, A. et al. Early boost and slow consolidation in motor skill learning. Learn Mem.;2006;13(5):580–3.doi:10.1101/lm.239406

70. Poldrack RA, Sabb FW, Foerde K, et al. The neural correlates of motor skill automaticity. J Neurosci. 2005;25(22):5356–5364. doi:10.1523/JNEUR0SCI.3880-04.2005

71. Yang, J. The influence of motor expertise on the brain activity of motor task performance: A meta-analysis of functional magnetic resonance imaging studies. Cogn Affect Behav Neurosci. 2015; 15(2):381–94. doi: 10.3758/s13415-014-0329-0.

72. Schneider CL, Majewska AK, Busza A. et al. Selective serotonin reuptake inhibitors for functional recovery after stroke: Similarities with the critical period and the role of experience-dependent plasticity. J Neurol. 2019;1–7. doi:10.1007/s00415-019-09480-0

73. Stagg, CJ., Bachtiar, V., Johansen-Berg, H. The role of GABA in human motor learning. Curr Biol. 2011; 21(6):480–4. doi: 10.1016/j.cub.2011.01.069.

74. Stagg CJ, Bachtiar V, Amadi U, et al. Local GABA concentration is related to network-level resting functional connectivity. Elife. 2014;3:e01465. doi:10.7554/eLife.01465

75. Chen, Z., Silva, AC., Yang, J. et al. Elevated endogenous GABA level correlates with decreased fMRI signals in the rat brain during acute inhibition of GABA transaminase. J Neurosci Res. 2005; 79(3):383–91.

76. Bhagwagar, Z., Wylezinska, M., Taylor, M. et al. Increased brain GABA concentrations following acute administration of a selective serotonin reuptake inhibitor. Am J Psychiatry. 2004; 161(2):368–70. doi: 10.1176/appi.ajp.161.2.368

77. El Mansari, M., Sánchez, C., Chouvet, G. et al. Effects of acute and long-term administration of escitalopram and citalopram on serotonin neurotransmission: an in vivo electrophysiological study in rat brain. Neuropsychopharmacology. 2005; 30(7):1269–77. doi:10.1038/sj.npp.1300686

78. Lladó-Pelfort, L., Santana, N., Ghisi, V. et al. 5-HT1A receptor agonists enhance pyramidal cell firing in prefrontal cortex through a preferential action on GABA interneurons. Cereb Cortex:. 2012; 22(7):1487–97. doi: 10.1093/cercor/bhr220.

79. Karim HT, Huppert TJ, Erickson KI, et al. Motor sequence learning-induced neural efficiency in functional brain connectivity. Behav Brain Res. 2017;319:87–95. doi:10.1016/j.bbr.2016.11.021

80. Mushiake, H., Inase, M., Tanji, J. Neuronal activity in the primate premotor, supplementary, and precentral motor cortex during visually guided and internally determined sequential movements. J Neurophysiol; 1991;66(3):705–18.

81. Halsband, U., Ito, N., Tanji, J. et al. The role of premotor cortex and the supplementary motor area in the temporal control of movement in man. Brain; 1993;116 (Pt1):243–66. doi:10.1093/brain/116.1.243.

82. Kalaska, JF., Crammond, DJ. Deciding not to GO: neuronal correlates of response selection in a GO/NOGO task in primate premotor and parietal cortex. Cereb Cortex.; 1995; 5(5):410–28. doi:10.1093/cercor/5.5.410.

83. Bianchini F, Verde P, Colangeli S, et al. Effects of oral contraceptives and natural menstrual cycling on environmental learning. BMC Womens Health. 2018;18(1):179. Published 2018 Nov 7. doi:10.1186/s12905-018-0671-4

84. Will TR, Proaño SB, Thomas AM, et al. Problems and Progress regarding Sex Bias and Omission in Neuroscience Research. eNeuro. 2017;4(6):ENEURO.0278-17.2017. Published 2017 Nov 9. doi:10.1523/ENEURO.0278-17.2017

85. Henry ME, Lauriat TL, Lowen SB, et al. Effects of citalopram and escitalopram on fMRI response to affective stimuli in healthy volunteers selected by serotonin transporter genotype. Psychiatry Res. 2013;213(3):217–224. doi:10.1016/j.pscychresns.2013.05.008

86. Finnema SJ, Nabulsi NB, Eid T, et al. Imaging synaptic density in the living human brain. Sci Transl Med. 2016;8(348):348ra96. doi:10.1126/scitranslmed.aaf6667

87. Chen MK, Mecca AP, Naganawa M, et al. Assessing Synaptic Density in Alzheimer Disease With Synaptic Vesicle Glycoprotein 2A Positron Emission Tomographic Imaging. JAMA Neurol. 2018;75(10):1215–1224. doi:10.1001/jamaneurol.2018.1836

